# The role of population structure when measuring COVID-19 impact across countries

**DOI:** 10.1101/2020.11.30.20239947

**Authors:** Octavio Bramajo, Mauro Infantino, Rafael Unda, Walter D Cardona-Maya, Pablo Richly

## Abstract

The search for accurate indicators to compare the pandemic impact between countries is still a challenge. The crude death rate, case fatality rate by country and sex, standardized fatality rate, and standardized death rate were calculated using data from Argentina and Colombia countries. We show that even when frequently used indicator as deaths per million are quite similar, 512 deaths per million in Argentina and 522 deaths per million in Colombia, a significant heterogeneity can be found when the mortality data is decomposed by sex or age.

## Introduction

One of the most significant challenges has been determining the most accurate indicators to compare the pandemic impact between different regions (e.g. countries). There is no doubt that the pandemic of the coronavirus disease 2019 (COVID-19) has had great health and economic impact globally, although it is still difficult to measure it accurately. It is evident that its effect has been heterogeneous in the different continents, and there is a consensus that it has been less harmful in Asia, Africa, and Oceania than in Europe or America.

The search for additional indicators to illustrate both the recent and accumulated effect of infections has popularized internet pages such as Our World In Data (1) or Worldometer (2). Within these indicators, the number of deaths per million inhabitants turned out to be one of the preferred ones to generate a ranking of the most affected countries.

These indicators’ interpretation could be conditioned by variables such as demographic structure, socioeconomic status, or available health system resources.

In order to analyze these possible differences, we have scoped to draft a mortality profile of two South American countries that share, in addition to the same geographic region, other similarities such as the number of inhabitants: Argentina and Colombia.

On March 2, 2020, the first patient was detected in Argentina. Meanwhile, on March 6, the first case of COVID-19 was confirmed in Colombia. Nine months after the first infections, after more than 52.6 million infections worldwide and more than 1.29 million deaths worldwide. Specifically, Argentina and Colombia countries report similar figures for infection: 1.28 and 1.17 million cases, and 34,782 deaths in Argentina and 33,491 in Colombia.

Therefore, this study aimed to explore the series of possible heterogeneities that could be found beneath the apparent similarity between two countries that appear to be matched when compared by indicators such as mortality.

## Materials and methods

We performed an ecological type study using aggregate data to explore differentials at a population level. To do so, we took advantage of the daily report freely available of the Health Ministry of Argentina (MSAL) - http://datos.salud.gob.ar/dataset/covid-19-casos-registrados-en-la-republica-argentina-, with data provided via the Integrated System of Health Information and Colombian data supplied by the National Health Institute (Colombia) - https://www.ins.gov.co/Noticias/Paginas/Coronavirus.aspx-. The data analyzed belongs to the period January 1^st^ -September 30^th^ and was extracted on October 31^st^ to minimize missing information due to delays in the data registration.

Given that the data were anonymized, public, and freely available, informed consents were not needed nor ethical committee approval. The data were taken from the public domain, which may not be accurate or confirmed by nations’ public health units.

In the epidemiological analysis of COVID-19, the case fatality ratio (CFR) was a standard measure of the intensity of the pandemic. CFR is essentially the quotient between Deaths (D) and positive cases (C) detected.

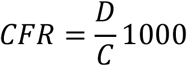

However, there has been less emphasis in death rates attributed to COVID-19. This is mostly due to the lack of a full year of exposure, given that death rates are calculated (or at least they should be) as the quotient of deaths (D) and the mean population (N) in a given year (conventionally expressed in the population at June 30 or July 1). Actually, the mean population represents the average time that a given population has lived in a year, expressed in person-years as a result (3). However, since from March to September there is only a seven-year period of exposure (and not an entire year), we could opt for either a) assume the same mortality intensity by modifying the exposure for a proportional value to calculate a crude death rate b) calculating a partial crude death rate (PCDR), expressed as:

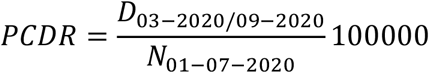

In our vision, this is the least problematic way to calculate death rates. The first route relies in some robust assumptions that maybe not be justified, given that seasonality may play a part in COVID-19 mortality.

When information about death distributed by age groups is available, both CFR and PCDR could be expressed as the sum of the different death proportions (P) of cases (C) and population (N) respectively by age groups (e), which also could help us calculating rates in a given age group or interval.

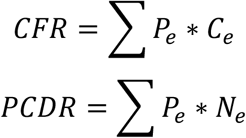

However, given that population structure usually plays a part in calculating death rates (4) and fatality ratios (5), calculating standardized rates, by using the corresponding weight of the overall sum of positive cases detected (CW) and the weight of population (NW) of both countries respectively by ten age-groups, is desirable to make a more precise comparison. Standardized case fatality ratios (SCFR) and standardized partial death rates (SPDR) should be expressed as the following.

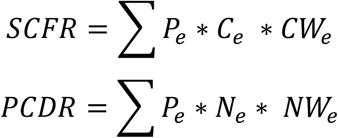

After some preliminary testing, we observed that a small number of positive cases (less than 1%) in Argentina had no province of residence reported. Those were excluded from the analysis. Furthermore, 1486 deaths (6,4% of total deaths) had jurisdiction of residence assigned but not sex. Hence, sex was imputed by using a simple proportional criterion (3). Finally, we only considered cases, population, and deaths of individuals aged between 30 and 99 years to avoid noises with centenarians and avoid overrepresenting the lower fatality in the younger ages in summary measures.

## Results

As of the database’s cut-off date (30^st^ September 2020), there are no essential differences regarding mortality and global fatality from COVID-19 between the two countries. The difference in fatality between Argentina and Colombia is 0.75 points per thousand.

In both countries, both mortality and fatality are higher in men than in women, although this difference is more noticeable in Colombia. Remarkably, both mortality and non-standardized fatality rates in women seem to be higher in Argentina, while it is higher in men in Colombia. The mortality difference between Argentina and Colombia is 8.37 points per thousand in women and 22.55 points per thousand in men, while the mortality difference is 36.70 and 66.97 per hundred thousand points, respectively.

If we calculate the percentage of deaths under 70 years of age, age structures’ impact is evident in the total estimates. There is almost 15% more “premature” mortality in Colombia compared to Argentina in women, and little more than 5% in men. It is also worth mentioning how the gender gap in Argentina is notorious.

When the standardized fatality ratio (SFR) and standardized partial death rate (SPDR) are calculated, differences in fatality ratio for females tend to be less pronounced when compared to the CFR. Simultaneously, the gradient in the partial death rate reverses after standardization: Colombia presents the highest standardized partial death rate (SPDR), both for males and females. It must be noticed that, for both countries, the gap in CPDR between males tends to enlarge after standardization, but in terms of fatality ratio, it tends to decrease slightly.

## Discussion

In the present study, it was observed that Argentina and Colombia have a CDR of 51.6 and 52.2 in people over 30 and under 100 years old, respectively. Results similar analyzing data from 121 countries, 46 high-income, 36 upper-middle-income and 39 low, they found that the CDR of high countries is 2.16 times higher compared to upper-middle-income countries (166 vs. 77) and 6.1 times higher than poor countries (166 vs. 27) were recently reported (6).

A figure that suggests a similar impact of the pandemic (512 deaths per million in Argentina vs. 522 deaths per million in Colombia) is put to the test by carrying out a more detailed analysis of its specific effect concerning the composition of each one of the populations. While the data in Table 1 do not show any striking discrepancies in the mortality profile due to COVID-19 in the countries selected in our study, we find that only by decomposing the population with respect to sex the first differences emerge. However, the same differential cannot be directly extrapolated with respect to the pandemic’s adverse effects since they are not standardized by age.

**Table 1.** CFR and CDR by country.

| Country | Cases | Deaths | Population | CFR | CDR |
| --- | --- | --- | --- | --- | --- |
| Argentina | 820309 | 23326 | 45195777 | 28.4 | 51.6 |
| Colombia | 910513 | 26580 | 50882884 | 29.2 | 52.2 |

**Table 2.** CFR and CDR by country and sex.

| Country | Sex | Cases | Deaths | Population | CFR | CDR |
| --- | --- | --- | --- | --- | --- | --- |
| Argentina | Female | 285180 | 10037 | 12523929 | 35.2 | 80.1 |
| Argentina | Male | 297342 | 12954 | 11086967 | 43.6 | 117.0 |
| Colombia | Female | 304852 | 9323 | 13934364 | 30.6 | 66.9 |
| Colombia | Male | 316547 | 16820 | 12563425 | 53.1 | 134.0 |

**Table 3.** Premature death by country and sex.

| Country | Sex | Deaths <70 years | Total deaths | Percentage |
| --- | --- | --- | --- | --- |
| Argentina | Female | 2863 | 10037 | 28.5 |
| Argentina | Male | 5668 | 12954 | 43.8 |
| Colombia | Female | 3985 | 9323 | 42.7 |
| Colombia | Male | 8273 | 16820 | 49.2 |

**Table 4.** SFR and SPDR by country and sex.

| **Country** | **Sex** | **SFR (\*1000)** | **SDR (\*1000000)** |
| --- | --- | --- | --- |
| Argentina | Female | 31.4 | 63.0 |
| Argentina | Male | 48.6 | 124.7 |
| Colombia | Female | 30.6 | 69.3 |
| Colombia | Male | 53.2 | 154.6 |

When we consider age composition, some dimensions of the impact of the pandemic between the two countries become much more evident, as well as the previously observed differences with respect to sex in context, showing that higher rates would correspond to a more aged population.

It should be considered that these analyzes have only data from the population between 31 and 99 years of age to avoid the distorting effect of the few deaths at ages outside this range. However, we consider that the present work eloquently shows that a global analysis of mortality is not sufficient to apprehend the specific effect of the pandemic in each country, even in cases where one can assume that they are comparable.

The limitations of the present study and suggestions for future research are addressed. To compare deaths and cases by sex and age in a given date, we decided to consider data reported until September 30, 2020. In such records, we observed a series of sociodemographic aspects (like sex, age, and region of residence) of individuals reported as suspect cases of COVID-19 and the moment of detection of the disease in those who turned out to be positive cases. Additionally, they also record the date of death. However, it must be mentioned that such information is sensitive to manual data processing errors, and that *ex post* health authorities may submit the records to modifications and changes. That means, even if those data are updated continuously, they are not a replacement for the vital statistics provided by each country.

Furthermore, not every death assigned to a positive case implies that COVID-19 was the cause of death. The present study’s objective is not to establish the “real” impact of mortality attributed to COVID-19 in Argentina and Colombia, but make an approximation given on the know information and the available sources.

In conclusion, any death rate comparison that is not sex or age adjusted might lead to a misrepresentation of the actual differences of the pandemic impact between countries.

## Data Availability

All the data analyzed can be is available at the links listed below.

http://datos.salud.gob.ar/dataset/covid-19-casos-registrados-en-la-republica-argentina

https://www.ins.gov.co/Noticias/Paginas/Coronavirus.aspx

